# Rapid emergence of SARS-CoV-2 Omicron variant is associated with an infection advantage over Delta in vaccinated persons

**DOI:** 10.1101/2022.01.22.22269660

**Authors:** Chrispin Chaguza, Andreas Coppi, Rebecca Earnest, David Ferguson, Nicholas Kerantzas, Frederick Warner, H. Patrick Young, Mallery I. Breban, Kendall Billig, Robert Tobias Koch, Kien Pham, Chaney C. Kalinich, Isabel M. Ott, Joseph R. Fauver, Anne M. Hahn, Irina R. Tikhonova, Christopher Castaldi, Bony De Kumar, Christian M. Pettker, Joshua L. Warren, Daniel M. Weinberger, Marie L. Landry, David R. Peaper, Wade Schulz, Chantal B.F. Vogels, Nathan D. Grubaugh

## Abstract

The emergence of severe acute respiratory syndrome coronavirus 2 (SARS-CoV-2) variants continues to shape the coronavirus disease 2019 (Covid-19) pandemic. The detection and rapid spread of the SARS-CoV-2 ‘ Omicron’ variant (lineage B.1.1.529) in Botswana and South Africa became a global concern because it contained 15 mutations in the spike protein immunogenic receptor binding domain and was less neutralized by sera derived from vaccinees compared to the previously dominant Delta variant. To investigate if Omicron is more likely than Delta to cause infections in vaccinated persons, we analyzed 37,877 nasal swab PCR tests conducted from 12-26 December 2021 and calculated the test positivity rates for each variant by vaccination status. We found that the positivity rate among unvaccinated persons was higher for Delta (5.2%) than Omicron (4.5%). We found similar results in persons who received a single vaccine dose. Conversely, our results show that Omicron had higher positivity rates than Delta among those who received two doses within five months (Omicron = 4.7% vs. Delta = 2.6%), two doses more than five months ago (4.2% vs. 2.9%), and three vaccine doses (2.2% vs. 0.9%). Our estimates of Omicron positivity rates in persons receiving one or two vaccine doses were not significantly lower than unvaccinated persons but were 49.7% lower after three doses. In comparison, the reduction in Delta positivity rates from unvaccinated to 2 vaccine doses was 45.6-49.6% and to 3 vaccine doses was 83.2%. Despite the higher positivity rates for Omicron in vaccinated persons, we still found that 91.2% of the Omicron infections in our study occurred in persons who were eligible for 1 or more vaccine doses at the time of PCR testing. In conclusion, escape from vaccine-induced immunity likely contributed to the rapid rise in Omicron infections.

## Main text

The emergence of severe acute respiratory syndrome coronavirus 2 (SARS-CoV-2) variants continues to shape the coronavirus disease 2019 (Covid-19) pandemic^1^. Mathematical modeling suggests that SARS-CoV-2 variants with increased transmissibility and partial immune escape may significantly increase infections even in a well-immunized population^2^. The detection and rapid spread of the SARS-CoV-2 ‘ Omicron’ variant (lineage B.1.1.529) in Botswana and South Africa was thus a global concern because it contained 15 mutations in the spike protein immunogenic receptor binding domain^3,4^. Subsequent *in vitro* assays showed that antibody-mediated neutralization using sera derived from vaccinees was significantly lower for Omicron than the previously dominant Delta variant^5–8^. For example, comparing Omicron to Delta, antibody neutralization was diminished 17x following two doses of mRNA vaccine (>28 days post vaccination)^9^ and 25x following three doses of BTN162b2 (>3 months post vaccination)^10^. While these data suggest that Omicron may have an infection advantage over Delta in vaccinated persons, *in vitro* neutralization is not a direct correlate for human protection from infection.

We established a surveillance program in southern Connecticut, USA to investigate the emergence of Omicron using a combination of spike-gene target failure (SGTF) polymerase chain reaction (PCR) signatures and sequencing. We first detected Omicron (lineage BA.1) on 4 December 2021, and it became the dominant variant in our cohort on 20 December, 16 days after first detection (**Figure 1a**). Fitting an exponential curve to cumulative cases, we estimated that Omicron cases doubled every 3.2 days (95% confidence interval (CI): 3.0-3.5), 3.7x shorter than the initial doubling time for Delta during its emergence period from 18 April to 26 May, 2021 (11.9 days [95% CI: 10.7-13.3]; **Figure S1**). When we first detected Omicron, 71-74% of the population in our testing catchment area had completed a primary COVID-19 vaccine series (1 dose of Ad26.COV2.S or 2 doses of mRNA-1273 or BNT162b2)^11^. We hypothesized that part of the rapid increase in Omicron infections stemmed from its increased ability to cause infections in persons that are vaccinated compared with Delta.

**Figure 1.**
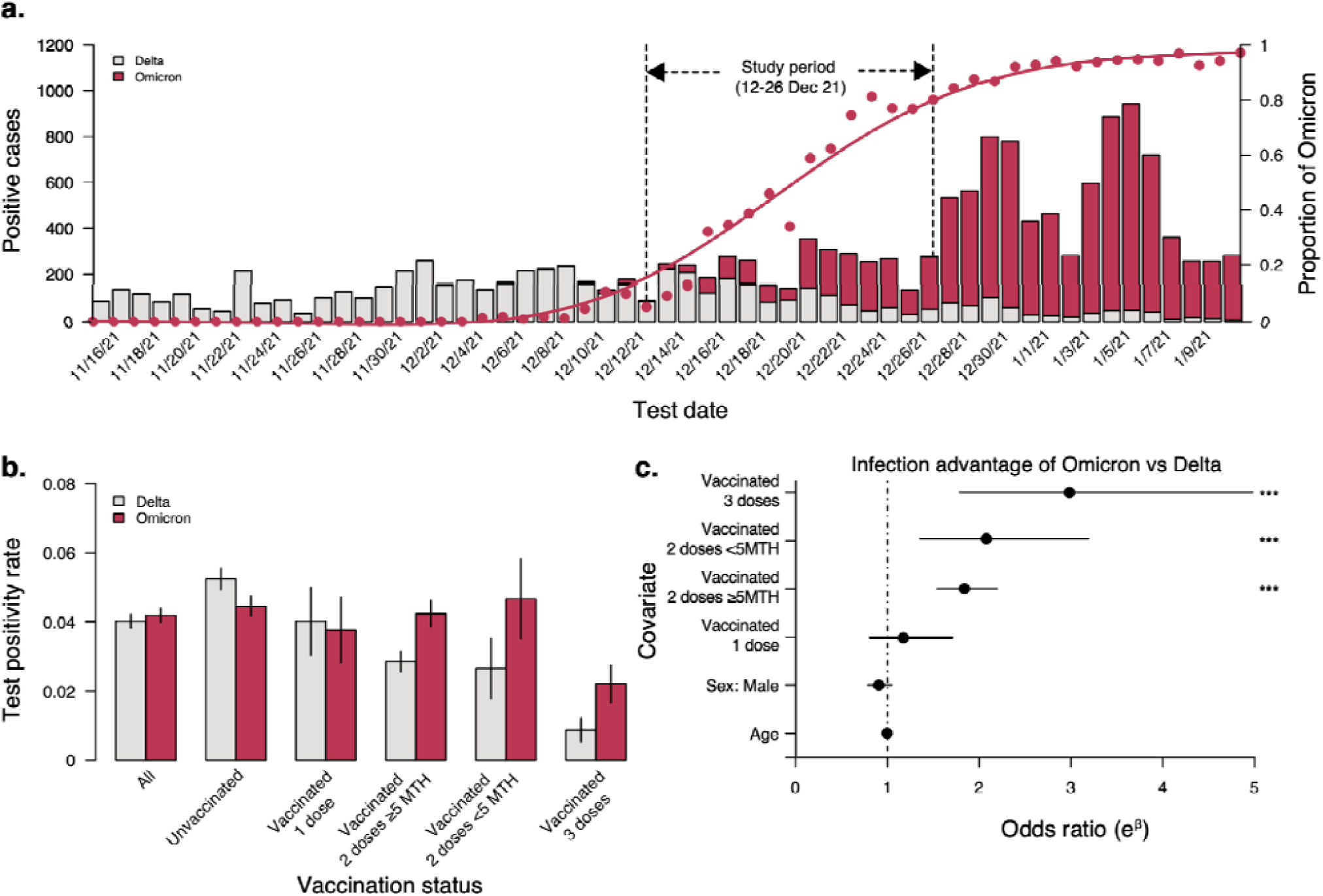
Variant case counts, test positivity, and odds of infection by vaccination status. **a)** Number of persons infected with Delta and Omicron SARS-CoV-2 variants and the proportion of Omicron cases in southern Connecticut. Overlayed on the plot showing the number of positive cases are the proportion of Omicron variants (dots) with a fitted smoothed curve. The growth rate of Omicron compared to Delta during their respective emergence periods is shown in **Figure S1. b)** The proportion of positive SARS-CoV-2 PCR tests (Ct < 30) for Delta and Omicron variants (using SGTF to differentiat) by vaccination status. The positivity rate values are listed in **Table S2. c)** Odds of infection with Omicron relative to Delta variants by age, sex and vaccination status among individuals who tested positive for SARS-CoV-2. We regressed the binary outcome for the SARS-CoV-2 variant (Delta as the reference group) and specified females and unvaccinated persons as the reference categories for the sex and vaccination status predictor variables in the model. Odds ratios > 1 indicate higher odds of detecting Omicron relative to Delta in persons testing positive for SARS-CoV-2 infection. The odds ratio values are listed in **Table S3**. The positivity rates and odds ratios stratified by vaccine manufacturers are shown in **Figure S2** and **Table S4**.

To investigate if Omicron is more likely than Delta to cause infections in vaccinated persons, we analyzed 37,877 nasal swab PCR tests conducted from 12-26 December when the total number of probable Delta and Omicron infections were relatively equal (Delta = 1463/2987, 49.0%; Omicron = 1524/2987, 51.0%). For each PCR test, we collected the date and manufacturer of each COVID-19 vaccine administered, and cycle threshold (Ct) values < 30 were differentiated as Omicron or Delta by SGTF signatures. Details regarding the characteristics of the population are provided in **Table S1**. We then calculated the test positivity rates for each variant by vaccination status (**Figure 1b, Table S2**). We found that the positivity rate among unvaccinated persons was higher for Delta (5.2% [95% CI: 4.9-5.6%]) than Omicron (4.5 [95% CI: 4.2-4.7%]). We found similar results in persons who received a single vaccine dose. Conversely, our results show that Omicron had higher positivity rates than Delta among those who received two doses within five months (Omicron = 4.7% [95% CI: 3.5-5.8%] vs. Delta = 2.6% [95% CI: 1.8-3.5%]), two doses more than five months ago (4.2% [95% CI: 3.9-4.6%] vs. 2.9% [95% CI: 2.5-3.2%]), and three vaccine doses (2.2% [95% CI: 1.7-2.7%] vs. 0.9% [95% CI: 0.5-1.2%]. Our estimates of Omicron positivity rates in persons receiving one or two vaccine doses were not significantly lower than unvaccinated persons but were 49.7% lower after three doses. In comparison, the reduction in Delta positivity rates from unvaccinated to 2 vaccine doses was 45.6-49.6% and to 3 vaccine doses was 83.2% (**Table S2**). Despite the higher positivity rates for Omicron in vaccinated persons, we still found that 91.2% (1401/1524) of the Omicron infections in our study occurred in persons who were eligible for 1 or more vaccine doses at the time of PCR testing.

We confirmed our test positivity analysis by calculating the odds of detecting Omicron relative to Delta using a logistic generalized estimating equation model (**Figure 1c, Table S3, Table S4**). For infections among persons who were vaccinated, we found higher odds that they were infected with Omicron (versus Delta), and that the odds increased with increased number of vaccine doses. The odds of infection did not vary by sex or age and our results were similar when we stratified the data by vaccine-manufacturer (**Figure S2**). These findings support our hypothesis that Omicron has an infection advantage in vaccinated persons relative to Delta.

As an additional analysis, we compared nasal swab PCR Ct values by vaccination status and variants. After adjusting for covariates, we found that the Ct values were mostly consistent across vaccine doses, but Omicron infections had higher Ct values (i.e., lower viral load) than those infected with Delta (**Figure S3, Table S5, Table S6**).

Our study had several limitations. First, we did not sequence all samples inferred by SGTF, although Omicron was found in all the representative sequenced samples. Second, we excluded from analysis persons with incomplete vaccine information. However, excluding these samples did not significantly decrease the sample size. Third, we did not have access to information regarding previous infections, so we could not study the impact on reinfections. Finally, our study focused on SARS-CoV-2 infections in persons getting tested (for a variety of indications) and not disease; hence, our findings do not imply that Omicron is more likely to cause disease in vaccinated persons than Delta.

In conclusion, escape from vaccine-induced immunity likely contributed to the rapid rise in Omicron infections. Our findings may also explain why Omicron has been associated with more reinfections^12^. While Omicron was more likely to cause infections in vaccinated persons than Delta, vaccination shows maintained effectiveness in reducing severe disease, even for Omicron^13^. Together with the rebound of vaccine effectiveness after administering a booster dose^14^, measures to expand the uptake of the primary vaccine series and additional booster doses remain an important strategy for controlling the COVID-19 pandemic.

## Data Availability

All data produced in the present study are available upon reasonable request to the authors

## Acknowledgements

We would like to thank the Yale New Haven Health COVID-19 testing enterprise for collecting and testing samples and all of the health care workers supporting patients during the “Omicron surge”. This work was supported by CTSA Grant Number TL1 TR001864 (R.E.), Fast Grant from Emergent Ventures at the Mercatus Center at George Mason University (N.D.G.), and the Centers for Disease Control and Prevention (CDC) Broad Agency Announcement # 75D30120C09570 (N.D.G.).

## Supplementary Methods

### Study design

We established a surveillance program at Yale University in conjunction with Yale New Haven Health (YNHH) to investigate the local emergence of Omicron. Similar to the Alpha variant, most Omicron viruses have a spike gene deletion (Δ69/70 HV) that causes “spike gene target failure” (SGTF) when using the ThermoFisher TaqPath COVID-19 Combo Kit qRT-PCR assay, allowing us to quickly identify potential Omicron infections. Our SGTF case definition included having an ORF1ab gene target PCR cycle threshold (Ct) of < 30 and spike gene target “not detected”. YNHH uses TaqPath for testing anterior nares swabs from symptomatic and asymptomatic outpatients for SARS-CoV-2 at collection sites in New London, New Haven, and Fairfield Counties, Connecticut. Here, we retrospectively applied the SGTF case definition to samples collected since 15 November 2021 and prospectively to 10 January 2022 (**Figure 1a**). We detected the first sample meeting our SGTF case definition on 4 December, which we sequence-confirmed as Omicron lineage BA.1. We sequenced a subset of samples collected from 22 November to 27 December (*n* = 695), and 100% (216/216) of the SGTF samples were confirmed as Omicron (BA.1) and 100% (479/479) of samples without SGTF (i.e., the spike gene was detected) were confirmed as Delta (B.1.617.2 or AY.x; **Extended Data File 1**). This established our SGTF, and spike gene target detected case definitions as adequate proxies for Omicron and Delta infections, respectively, during our study period.

We selected a period from 12-26 December 2021 when probable Delta and Omicron infections were relatively equal (Delta = 1463/2987, 49.0%; Omicron = 1524/2987, 51.0%). We conducted a medical records review to identify 37,877 TaqPath PCR tests from 34,980 unique persons conducted during that period, categorizing negative tests, positive tests with Ct values ≥ 30, and positive tests with Ct values < 30 differentiated into Omicron and Delta. For each person tested, we collected information on age and sex of the person, test date, test outcome (negative, positive ≥ 30 Ct, positive Delta, and positive Omicron), and date and manufacturer of each COVID-19 vaccine administered at least 14 days prior to the test date. We excluded persons who indicated in their records a preference to opt out of research.

### Study oversight

The Institutional Review Board from the Yale University Human Research Protection Program determined that obtaining de-identified test results linked to vaccination status and sequencing of de-identified remnant COVID-19 clinical samples obtained from clinical partners conducted in this study is not research involving human subjects (IRB Protocol ID: 2000031374).

### Study participants

Our study consisted of 34,980 unique persons that tested for SARS-CoV-2 (37,877 tests) from outpatient sites, including mass testing locations, in New London, New Haven, and Fairfield Counties, Connecticut. Provided indications for testing were being symptomatic for COVID-19, exposure to a known case of COVID-19, required testing (e.g. for work, school, or travel), and testing prior to undergoing an aerosol generating procedure. The participants included a diversity of ages from 0-5 to > 60, and 55% were female. We did not obtain information about race or ethnicity. The vaccinated persons received Ad26.COV2.S, mRNA-1273, and/or BNT162b2. Details regarding the characteristics of the population are provided in **Table S1**.

### Study outcomes

We quantified the positivity rates for the Omicron and Delta SARS-CoV-2 variants in our cohort, and estimated the odds ratios of detecting Delta in persons testing positive by sex, age, and vaccination status category. We also calculated the doubling times (in days) for the Omicron and Delta variants to understand their transmissibility. Finally, we assessed the association between the nasal swab PCR Ct value and sex, age, variant, and vaccination status category stratified by vaccine manufacturer.

### PCR testing for variant differentiation

Anterior nares swabs from outpatient collection sites were tested for SARS-CoV-2 by the YNHH COVID-19 and Clinical Virology Laboratories using the MagMAX viral/pathogen nucleic acid isolation kit and TaqPath COVID-19 Combo Kit. The TaqPath qRT-PCR assay reports Ct values from three SARS-CoV-2 gene targets: ORF1ab, spike, and nucleocapsid. ORF1ab with Ct values < 30 were investigated for spike gene detection. If the spike gene was detected, the sample was categorized as “probable Delta” and if the spike gene was not detected (i.e., SGTF), the sample was categorized as “probable Omicron”.

### Sequence confirmation of variants

Anterior nares swabs in viral transport media were received from SARS-CoV-2 infections from YNHH. Nucleic acid was extracted from 300 µL of the original sample using the MagMAX viral/pathogen nucleic acid isolation kit, eluting in 75 µl of the elution buffer. The extracted nucleic acid was again tested for SARS-CoV-2 RNA using a “research use only” (RUO) RT-qPCR assay^1^, which generates a SGTF result similar to the TaqPath assay. For rapid confirmation of the initial suspected Omicron samples with SGTF, we used the NEBNext ARTIC SARS-CoV-2 Companion Kit and sequenced pooled libraries on the Oxford Nanopore Technologies (ONT) MinION. The standard NEB protocol with PCR Bead Cleanup was slightly modified by using V4 or V4.1 primer pools for amplicon generation, by including an additional bead cleanup step (1:1 beads:sample) after the NEBNext end prep reaction, and by scaling up the barcode ligation reaction by using 16 µL of end-prepped DNA. Final pooled libraries were quantified using the Qubit High Sensitivity dsDNA kit, and the ONT SQK-LSK109 protocol was followed to prime and load the ONT MinION for sequencing. Samples were processed in sets of 14-46 samples with 2 negative controls. The RAMPART application developed by the ARTIC Network was used to monitor the sequencing run until sufficient coverage was reached (https://artic.network/ncov-2019/ncov2019-using-rampart.html)^2^. The ARTIC bioinformatics pipeline was used to generate consensus genomes with fast basecalling done by MinKNOW (https://artic.network/ncov-2019/ncov2019-bioinformatics-sop.html). A threshold of 20x coverage was used to call consensus genomes, and negative controls were confirmed to completely consist of Ns.

For routine sequencing of samples with nucleocapsid gene target Ct values ≤ 35, we used the Illumina COVIDSeq Test RUO version. The protocol was slightly modified by using V4 primers for amplicon generation, by lowering the annealing temperature of the amplicon generation step to 63° C, and by shortening the tagmentation step to 3 minutes. Final libraries were pooled and cleaned before quantification with the Qubit High Sensitivity dsDNA kit. The resulting libraries were sequenced using a 2×150 approach on an Illumina NovaSeq at the Yale Center for Genome Analysis. Each sequenced sample had at least 1 million reads. Samples were typically processed in sets of 93 or 94 with negative controls incorporated during the RNA extraction, cDNA synthesis, and amplicon generation steps. The reads were aligned to the Wuhan-Hu-1 reference genomes (GenBank: MN908937.3) using BWA-MEM v.0.7.15^3^. Adaptor sequences were trimmed, primer sequences were masked, and consensus bases were called with simple majority > 60% frequency using iVar v1.3.1^4^ and SAMtools v1.7^5^. An ambiguous ‘ N’ was used when fewer than 20 reads were present at a site. In all cases, negative controls were analyzed and confirmed to consist of at least 99% Ns. For both rapid and routine sequencing, Pangolin v.3.1.17^6^ was used to assign lineages^7^. Consensus genomes were submitted to GISAID and included in weekly updates on our website (https://covidtrackerct.com/).

### Variant growth rates

We first defined the emergence period for Omicron and Delta as the time since its first detection (and sequence-confirmed) from samples tested by YNHH. We first detected Omicron on December 4th, 2021, and first detected Delta on April 18th, 2021. Using the proportion of SGTF samples as a proxy for Omicron and the proportion of sequence-confirmed lineages for Delta^8^ from samples obtained by YNHH, we ran a logistic regression analysis for each variant separately, with the proportion of samples corresponding to a specific variant category as the outcome and the number of days since the first detection of the variant as the predictor. We used samples collected from April 18, 2021 to May 25, 2021 while data for Omicron was from December 4, 2021 to January 10, 2022. We plotted the smoothed fitted curves for the emergence periods with their 95% confidence intervals (**Figure S1**), which shows the probability of a given sequence belonging to a specific variant category over time. We estimated the doubling time by fitting an exponential curve to cumulative cases over time for each variant and dividing log(2) by the resulting coefficient.

### Positivity rates

The PCR positivity rates for each variant were estimated using the ORF1ab Ct values < 30 and SGTF signatures to define as Omicron or Delta. For this analysis, ORF1ab Ct values from 30-40 were included as “negatives” as we could not assign a variant category, and thus the variant-specific positivity rates that we show are not the true overall test positivity rates. As Omicron infections tend to have higher Ct values than Delta (**Figure S2**), our analysis may be biased against Omicron. We estimated the positivity rates for different SARS-CoV-2 variants as the proportion of persons testing positive during the study period with PCR Ct value <30 for the ORF1ab and S gene targets. We calculated the confidence intervals for the proportion based on the standard errors for the binomial distribution. We show each rate with the 95% CI.

### Odds of infection with Omicron relative to Delta

To assess the odds of detecting Omicron relative to Delta variant in infected persons, we fitted a logistic generalized estimating equation (GEE) with an unstructured correlation structure to determine the effect of the covariates, namely, sex, age, and vaccination status stratified by the vaccine manufacturer. We used the GEEs to account for the repeated measures to multiple testing of the same person during the study period. Similarly, we fitted a GEE with Gaussian estimation to assess the association between the ORF1ab PCR Ct value with covariates, namely, sex, age, and vaccination status stratified by the vaccine manufacturer. We specified females and unvaccinated persons as the reference categories for the sex and vaccination status covariates in the model.

---

**Figure S1.**
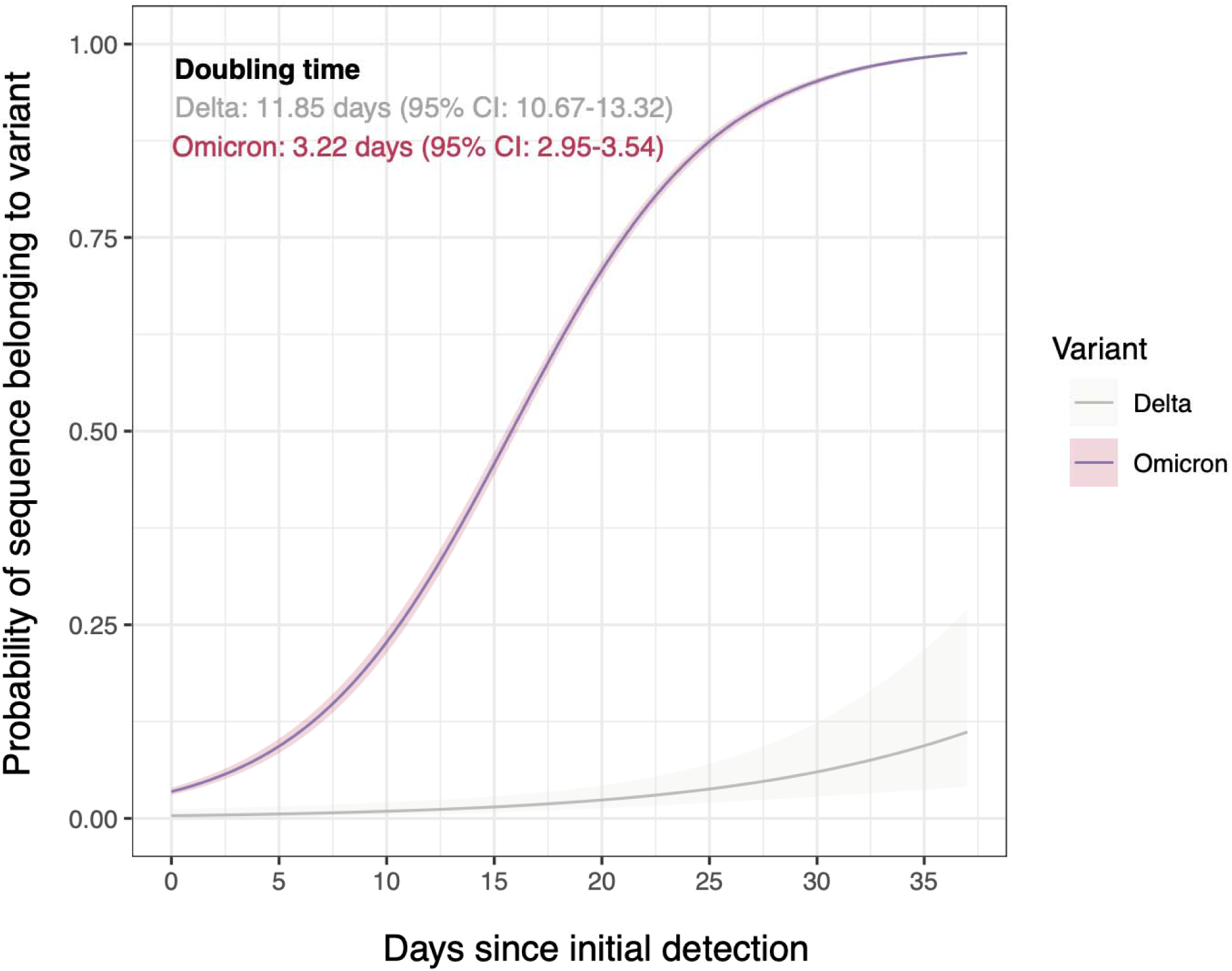
Growth rates of the Omicron and Delta SARS-CoV-2 variants during their emergence periods. Logistic regression analysis for each variant (run separately), showing the smoothed fitted curves of the probability of a given sequence belonging to a specific variant category for the emergence periods (38 days since first detection) with 95% confidence intervals. The curve for the growth dynamics of Delta was fitted for data collected from 18 April to 26 May, 2021 while for data for Omicron was from 4 December 2021 to 10 January 2022. Doubling times: Omicron 3.22 days (95% CI: 2.95-3.54); Delta 11.85 days (95% CI: 10.67-13.32).

**Figure S2.**
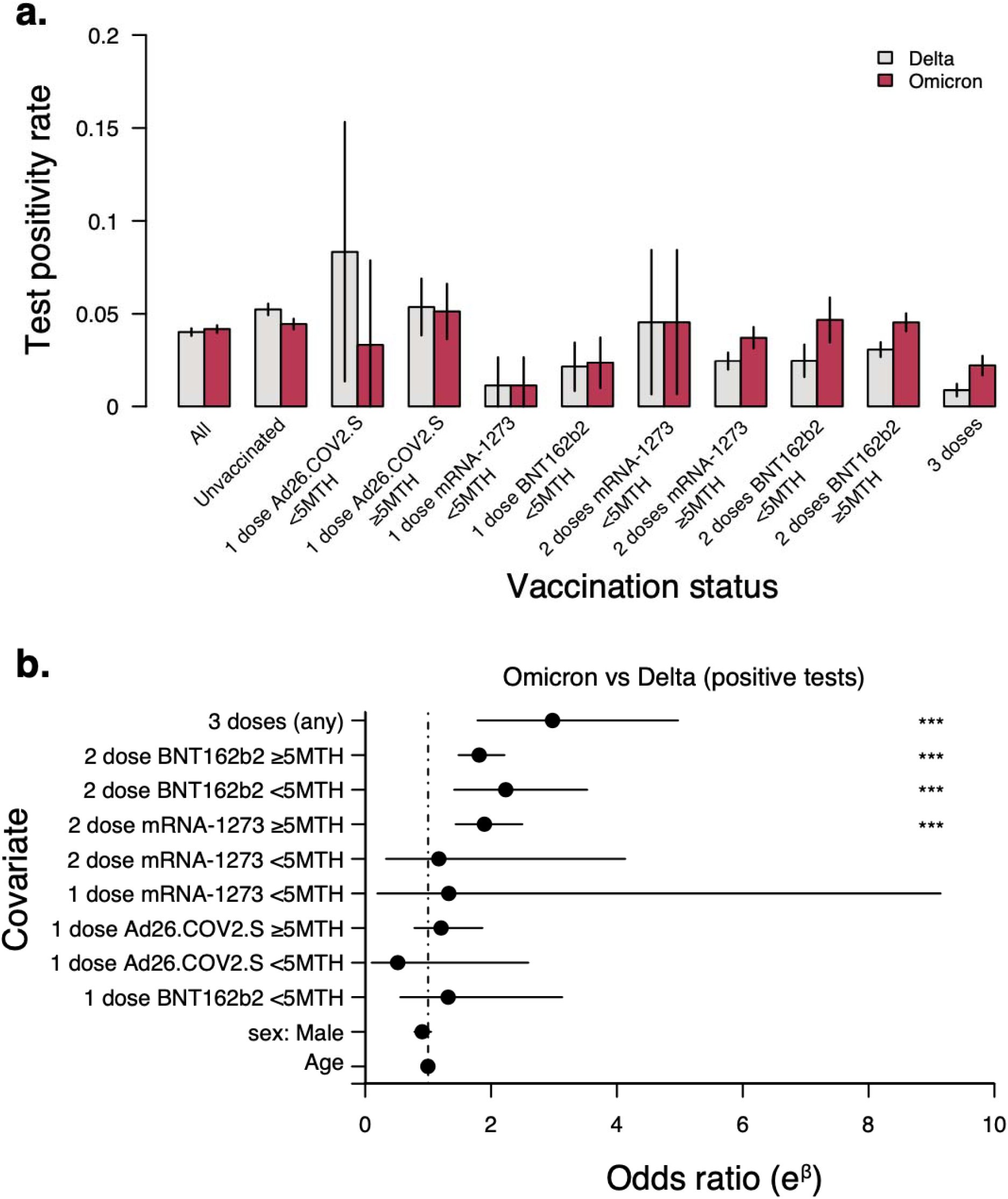
Test positivity, and odds of infection by vaccination status. **a)** The proportion of positive COVID tests for Delta and Omicron variants by vaccination status and vaccine manufacturer. **b)** Odds of infection with Omicron relative to Delta variants by age, sex, vaccination status, and vaccine manufacturer.

**Figure S3.**
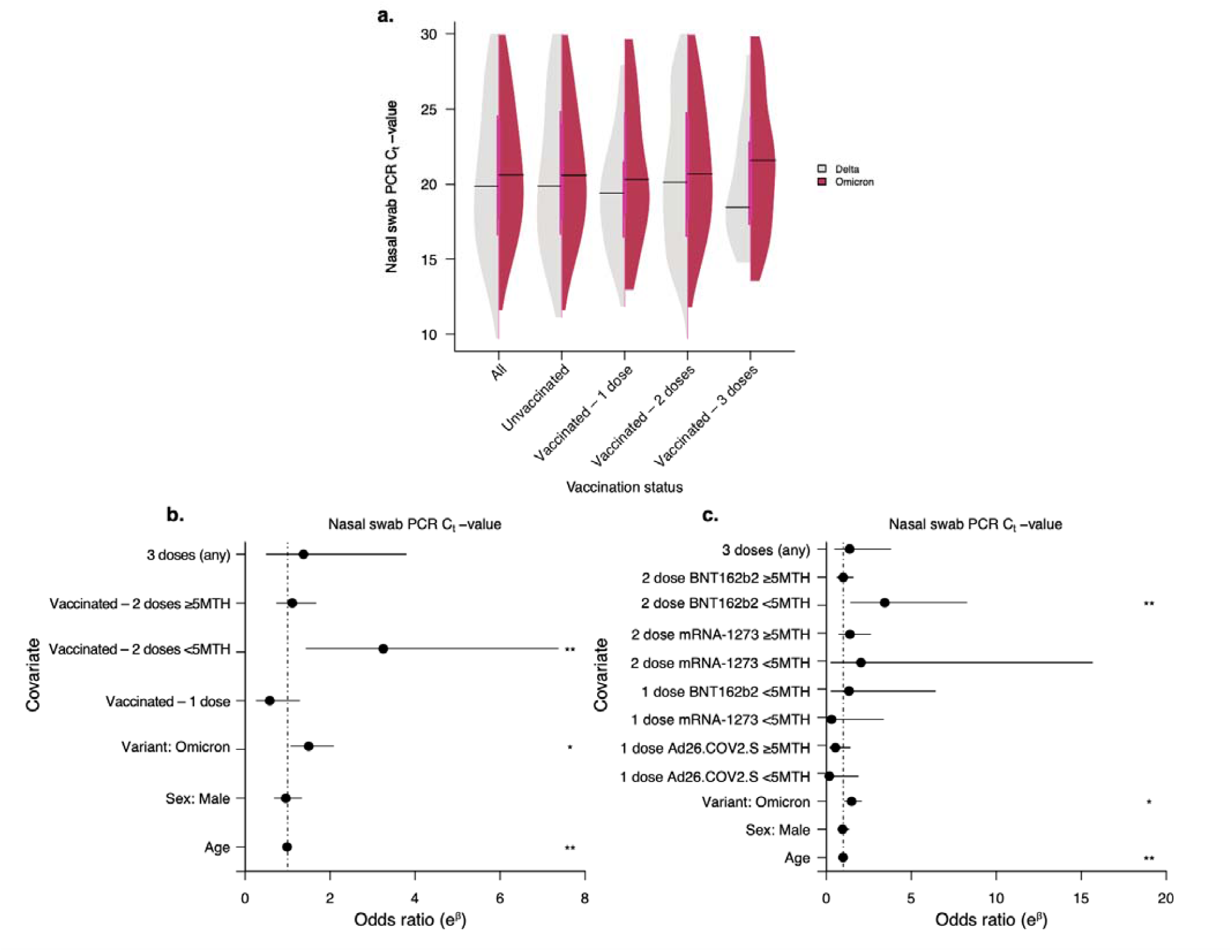
Effect of sex, age, variant and vaccination status on the nasal swab PCR cycle threshold (Ct). **a)** Nasal swab PCR Ct values for the Delta and Omicron SARS-CoV-2 variants by vaccination status. **b)** Association of age, sex, and vaccination status with PCR Ct values. Odds ratios > 1 indicate a higher CT value (lower virus RNA) for Omicron compared to Delta. The odds ratio values are shown in **Table S5. c)** Association of age, sex, vaccination status, and vaccine manufacturer with PCR Ct values. The odds ratio values are shown in **Table S6**.

**Table S1.** Demographic characteristics among PCR tests performed between 12-26 December 2021.

|  | Number of vaccine doses |  |  |  |
| --- | --- | --- | --- | --- |
| Age group, y | 0 (n=19522) | 1 (n=1543) | 2 (n=13955) | 3 (n=2857) |
| 0-5 | 3082 (15.79) | 33 (2.14) | 89 (0.64) | 0 (0) |
| 6-15 | 4139 (21.2) | 226 (14.65) | 1409 (10.1) | 1 (0.04) |
| 16-30 | 3811 (19.52) | 244 (15.81) | 2397 (17.18) | 262 (9.17) |
| 31-45 | 4282 (21.93) | 454 (29.42) | 3535 (25.33) | 737 (25.8) |
| 46-60 | 2652 (13.58) | 336 (21.78) | 3604 (25.83) | 803 (28.11) |
| >60 | 1556 (7.97) | 250 (16.2) | 2921 (20.93) | 1054 (36.89) |
| Sex |  |  |  |  |
| Female | 10731 (54.97) | 850 (55.09) | 8360 (59.91) | 1815 (63.53) |
| Male | 8778 (44.96) | 693 (44.91) | 5593 (40.08) | 1042 (36.47) |
| Unknown | 13 (0.07) | 0 (0) | 2 (0.01) | 0 (0) |

**Table S2.** Positivity rates for Omicron and Delta among PCR tests performed between 12-26 December 2021.

|  | Test positivity rate (Percentage, 95% CI) |  |  |  |  |  |
| --- | --- | --- | --- | --- | --- | --- |
| Variant | All (Vaccinated & unvaccinated, n=37877) | 0 dose – unvaccinated (n=19522) | Vaccinated – 1 dose (n=1543) | Vaccinated – 2 doses <5 months before test (n=1286) | Vaccinated – 2 doses ≥5 months before test (n=13955) | Vaccinated – 3 doses (n=2857) |
| Delta | 1463 (4.02: 3.82,4.22) | 1023 (5.24: 4.93,5.55) | 62 (4.02: 3.04,5) | 34 (2.64: 1.77,3.52) | 319 (2.85: 2.54,3.16) | 25 (0.88: 0.53,1.22) |
| Omicron | 1524 (4.19: 3.98,4.39) | 869 (4.45: 4.16,4.74) | 58 (3.76: 2.81,4.71) | 60 (4.67: 3.51,5.82) | 474 (4.24: 3.86,4.61) | 63 (2.21: 1.67,2.74) |
| All positive | 2987 (7.58: 7.32,7.85) | 1892 (8.84: 8.46,9.22) | 120 (7.22: 5.97,8.46) | 94 (6.81: 5.48,8.14) | 793 (6.62: 6.17,7.06) | 88 (2.99: 2.37,3.6) |

**Table S3.**
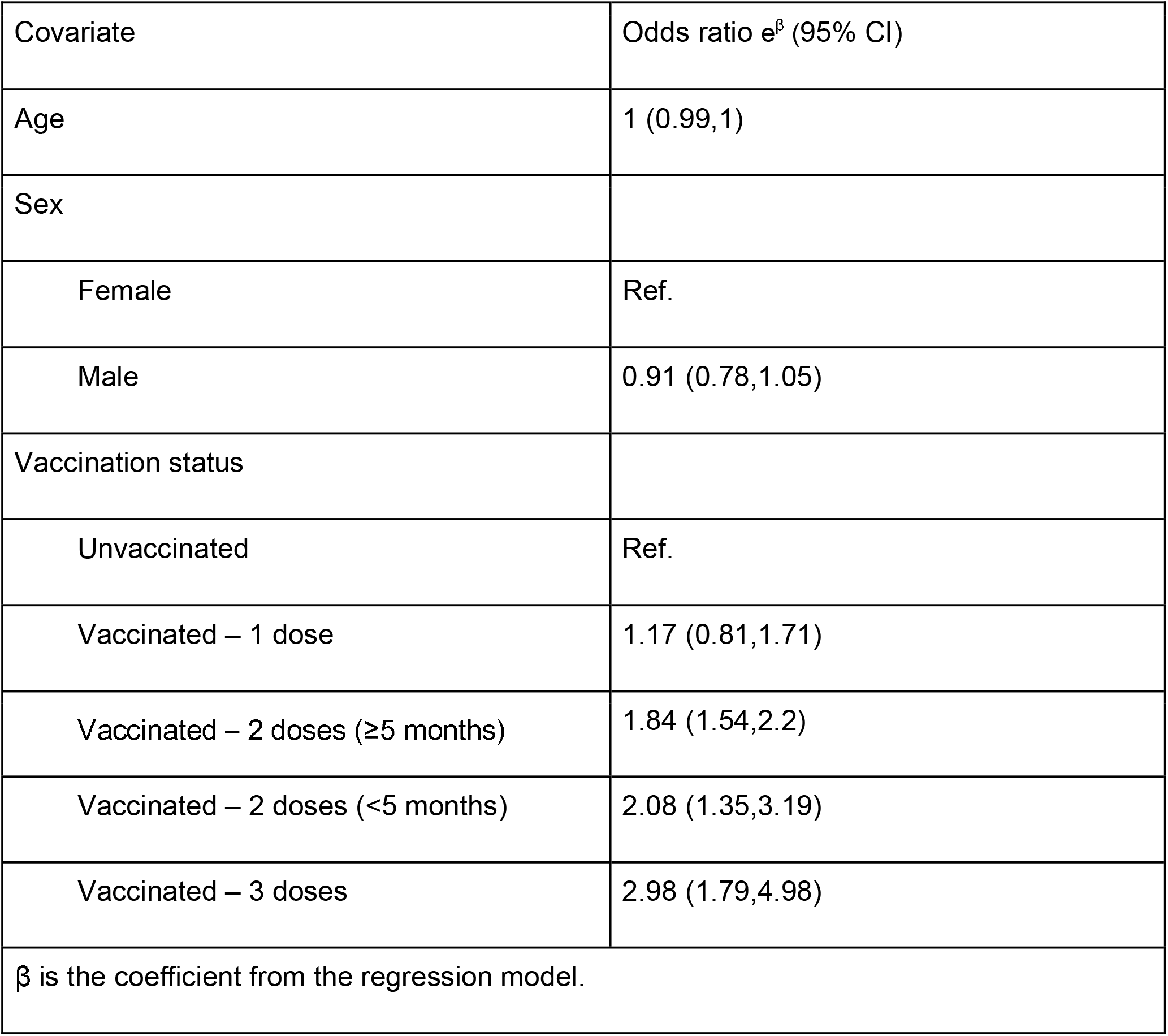
Odds of infection with Omicron relative to Delta SARS-CoV-2 variant by age, sex and vaccination status among positive PCR tests performed between 12-26 December 2021.

| Covariate | Odds ratio $e^{\beta}$ (95% CI) |
| --- | --- |
| Age | 1 (0.99,1) |
| Sex |  |
| Female | Ref. |
| Male | 0.91 (0.78,1.05) |
| Vaccination status |  |
| Unvaccinated | Ref. |
| Vaccinated – 1 dose | 1.17 (0.81,1.71) |
| Vaccinated – 2 doses ($\geq 5$ months) | 1.84 (1.54,2.2) |
| Vaccinated – 2 doses ($< 5$ months) | 2.08 (1.35,3.19) |
| Vaccinated – 3 doses | 2.98 (1.79,4.98) |
| $\beta$ is the coefficient from the regression model. | |

**Table S4.** Odds of infection with Omicron relative to Delta SARS-CoV-2 variant by age, sex, vaccination status, and vaccine manufacturer among positive PCR tests performed between 12-26 December 2021.

| Covariate | Odds ratio $e^{\beta}$ (95% CI) |
| --- | --- |
| Age | 1 (0.99,1) |
| Sex |  |
| Female | Ref. |
| Male | 0.91 (0.78,1.05) |
| Vaccination status |  |
| Unvaccinated | Ref. |
| 1 dose Ad26.COV2.S <5 months | 0.52 (0.1,2.59) |
| 1 dose Ad26.COV2.S ≥5 months | 1.21 (0.78,1.86) |
| 1 dose mRNA-1273 <5 months | 1.33 (0.19,9.15) |
| 2 dose mRNA-1273 <5 months | 1.17 (0.33,4.13) |
| 2 dose mRNA-1273 ≥5 months | 1.89 (1.44,2.5) |
| 1 dose BNT162b2 <5 months | 1.32 (0.55,3.13) |
| 2 dose BNT162b2 <5 months | 2.23 (1.41,3.53) |
| 2 dose BNT162b2 ≥5 months | 1.81 (1.48,2.21) |
| 3 doses (any) | 2.98 (1.78,4.97) |
| $\beta$ is the coefficient from the regression model. | |

**Table S5.** Association of the SARS-CoV-2 variant, age, sex and vaccination status with nasal swab PCR cycle threshold among positive PCR tests performed between 12-26 December 2021.

| Covariate | Effect size $e^{\beta}$ (95% CI) |
| --- | --- |
| Age | 1 (0.99,1) |
| Sex |  |
| Female | Ref. |
| Male | 0.91 (0.78,1.05) |
| Vaccination status |  |
| Unvaccinated | Ref. |
| 3 doses (any) | 2.98 (1.78,4.97) |
| 1 dose Ad26.COV2.S <5 months | 0.52 (0.1,2.59) |
| 1 dose Ad26.COV2.S ≥5 months | 1.21 (0.78,1.86) |
| 1 dose mRNA-1273 <5 months | 1.33 (0.19,9.15) |
| 2 dose mRNA-1273 <5 months | 1.17 (0.33,4.13) |
| 2 dose mRNA-1273 ≥5 months | 1.89 (1.44,2.5) |
| 1 dose BNT162b2 <5 months | 1.32 (0.55,3.13) |
| 2 dose BNT162b2 <5 months | 2.23 (1.41,3.53) |
| 2 dose BNT162b2 ≥5 months | 1.81 (1.48,2.21) |
| $\beta$ is the coefficient from the regression model. | |

**Table S6.**
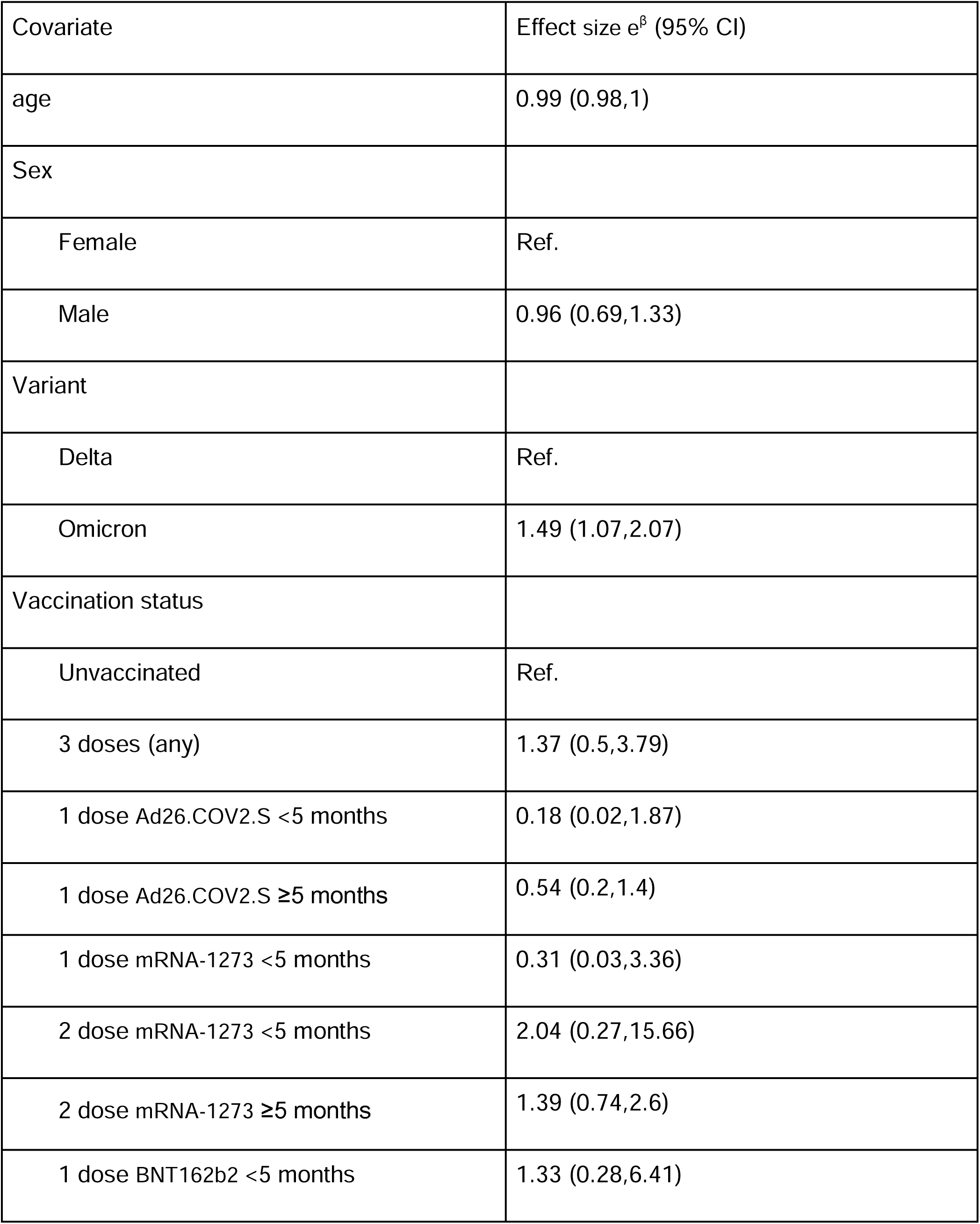
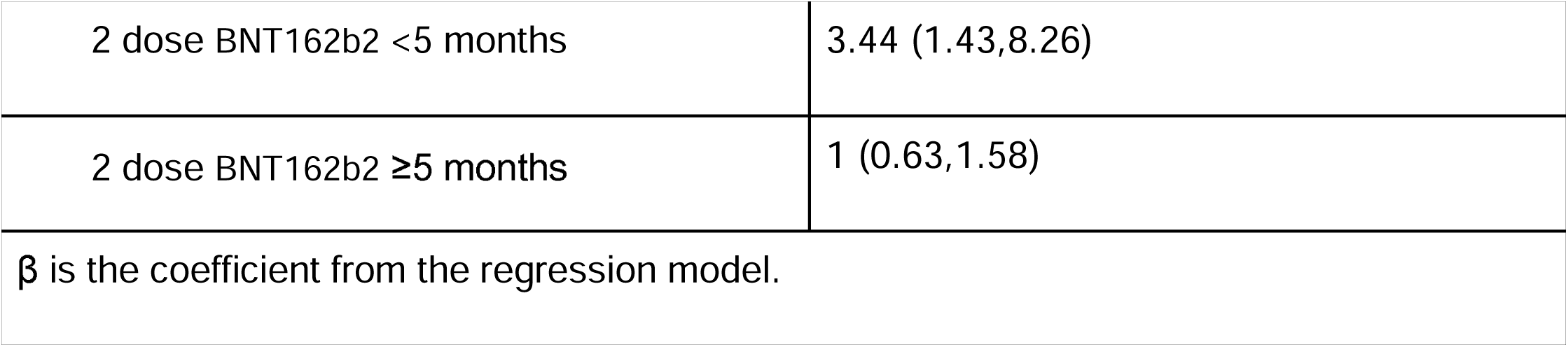
Association of the SARS-CoV-2 variant, age, sex, vaccination status, and vaccine manufacturer with nasal swab PCR cycle threshold among positive tests performed between 12-26 December 2021.

**Extended Data File 1: Validation of spike gene target failure (SGTF) as proxy for Omicron (BA.1) infection**. We compared results of our RUO RT-qPCR assay with sequencing results to show that SGTF is an adequate proxy for detection of Omicron (BA.1) in our study population. We sequenced a subset of samples collected from November 22nd to December 27th. Our N1 threshold was set at Ct <30.

